# Excess mortality and potential undercounting of COVID-19 deaths by demographic group in Ohio

**DOI:** 10.1101/2020.06.28.20141655

**Authors:** Troy Quast, Ross Andel

## Abstract

**Background:** There are significant gaps in our understanding of the mortality effects of COVID-19 due to evolving diagnosis criteria, shortages of testing supplies, and challenges faced by physicians in treating patients in crisis environments. Accurate information on the number of deaths caused by COVID-19 is vital for policy makers and health care providers.

**Methods:** We performed a retrospective study of weekly data for Ohio. To estimate expected mortality in 2020 we employed data from 2010 through 2019, adjusted for secular trends and seasonality. We estimated excess mortality as the number of observed deaths less the number of expected deaths. We conducted the analysis for the entire population and by age, gender, and county.

**Results:** We estimated 2,088 (95% CI 1,119-3,119) excess deaths due to natural causes in Ohio from March 15, 2020 through June 6, 2020. While the largest number excess of deaths was observed in the 80+ age group, our estimate of 366 (95% CI 110-655) excess deaths for those between 20 and 49 years of age substantially exceeds the reported number of COVID-19 deaths of 66.

**Conclusions:** Our methodology addressed some of the challenges of estimating the number of deaths caused by COVID-19. Our finding of excess deaths being considerably greater than the reported number of COVID-19 deaths for those aged 20 to 49 years old suggests that current tracking methods may not capture a significant number of COVID-19 deaths for this group. Further, increases in the infection rates for this cohort may have a greater mortality impact than anticipated.

## Introduction

A primary challenge in understanding COVID-19 has been the difficulty in accurately determining whether a death was caused by the novel coronavirus SARS-CoV-2. Evolving diagnosis criteria,^1^ testing supply constraints and the prioritization of tests for living patients,^2^ and the “fog of war” present in burdened intensive care units^3^ can pose obstacles to physicians in correctly identifying COVID-19 deaths. Accurate mortality data are crucial for effective medical and policy responses to the virus.

A potentially insightful approach to estimating COVID-19 deaths is to use historical data aggregated across multiple causes of death to estimate the expected (or counterfactual) number of deaths. The number of excess deaths due to the virus can then be estimated as the observed number of deaths less the expected number. This approach abstracts from errors due to the incorrect coding of deaths if the reported cause of death falls in the same grouping as the true cause of death. This methodology has been used to estimate deaths following natural disasters^4–7^ and deaths due to COVID-19.^8–15^ Prior COVID-19 analyses found varying levels of excess mortality, but largely focused on overall mortality with the exceptions of a study of Massachusetts that found little difference in excess mortality by gender^8^ and other studies that found excess deaths increased with age.^11, 13, 15^

The goal of this study was to estimate excess mortality in the context of the COVID-19 epidemic in Ohio from March 15, 2020 through June 6, 2020. Ohio is the seventh-largest U.S. state and a state that provides comprehensive health data online. In many ways the state is representative of the country, with a similar age profile and comparable median income.^16^ The state has been aggressive in controlling the COVID-19 outbreak by being the first state in the country to close its schools^17^ and soon thereafter closing restaurants and bars.^18^ The state began allowing businesses to reopen on May 4, 2020 and for the next eight weeks experienced falling rates of new cases and COVID-19 hospitalizations.^19^ We analyzed excess overall mortality as well as by gender, age, and county. We compared our excess mortality estimates to the number of reported COVID-19 deaths.

## Methods

### Study design and data sources

We performed a weekly time series analysis in which ten years of historical data were used to estimate the expected number of deaths in 2020. We defined excess mortality as the number of observed deaths less the expected number. Our mortality data were obtained from the Ohio Public Health Information Warehouse published by the Ohio Department of Health (ODH). The data for 2010-2018 were finalized, while data for 2019-2020 were provisional.

We obtained the number of reported COVID-19 deaths from the ODH’s COVID-19 dashboard. These data are reported by county health departments to the state via the Ohio Disease Reporting System. For each death the county of residence, gender, and age are reported, as well as the dates of COVID-19 onset, hospital admission, and death. The data from the Ohio Public Health Information Warehouse and ODU’s COVID-19 dashboard were obtained on July 14, 2020.

The overall population data employed were the 2018 vintage version of bridged-race population postcensal estimates published by the National Vital Statistics System branch of the U.S. Centers for Disease Control and Prevention. To approximate values for 2019 and 2020, for each subgroup we calculated the 2013-2018 compound annual growth rate and applied the rate to the 2018 value.

The study population was all Ohio residents from January 3, 2010 through June 6, 2020.

### Statistical analysis

Our baseline period was the full set of epidemiological weeks for each year from 2010 through 2019. The observation period begins with the week ending March 21, 2020, which was the week during which the first COVID-19 death was reported in Ohio, and ends with the week ending June 6, 2020. The observed number of deaths refers to the actual number of deaths in 2020, while the expected number of deaths corresponds to the estimate based on data from the baseline period. As the variation in the number deaths is greater than the mean, an over-dispersed log-linear model was used to estimate the expected number of deaths. The exposure variable was the relevant population. Annual indicator variables were included to account for secular trends in the number of deaths, while harmonic variables consisting of four Fourier terms were used to adjust for seasonality. The model was estimated separately for all residents and by gender, age group, and county. The estimating equation was:

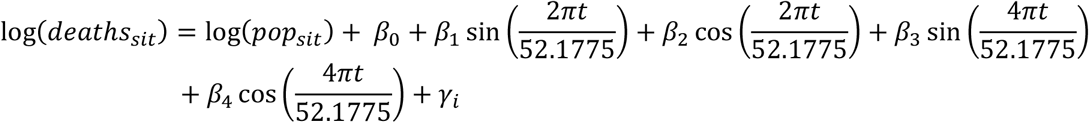

where *deaths* is the number of all-cause deaths for subgroup *s* in epidemiological year *i* and week *t*.

The variation in the expected number of deaths was modeled via parametric bootstrapping and follows an earlier study of COVID-19 excess mortality.^9^ From each regression we obtained the asymptotic covariance matrix, which was then used along with the estimated parameter values to specify a multivariate normal distribution to approximate the sampling distribution. One hundred samples were drawn from this distribution from which the mean number of deaths was calculated for each. We then drew one hundred samples from the Poisson distribution of each of these means, resulting in 10,000 samples for each subgroup and week. The 95% confidence interval bounds were estimated as the 2.5^th^ and 97.5^th^ percentile of the distribution. We also calculated the number of observed deaths less the number of reported COVID-19 deaths for each subgroup.

The weekly estimates are displayed Figures 1-4 while Appendix Table 2 contains the excess death estimates for the entire observation period.

**Figure 1.**
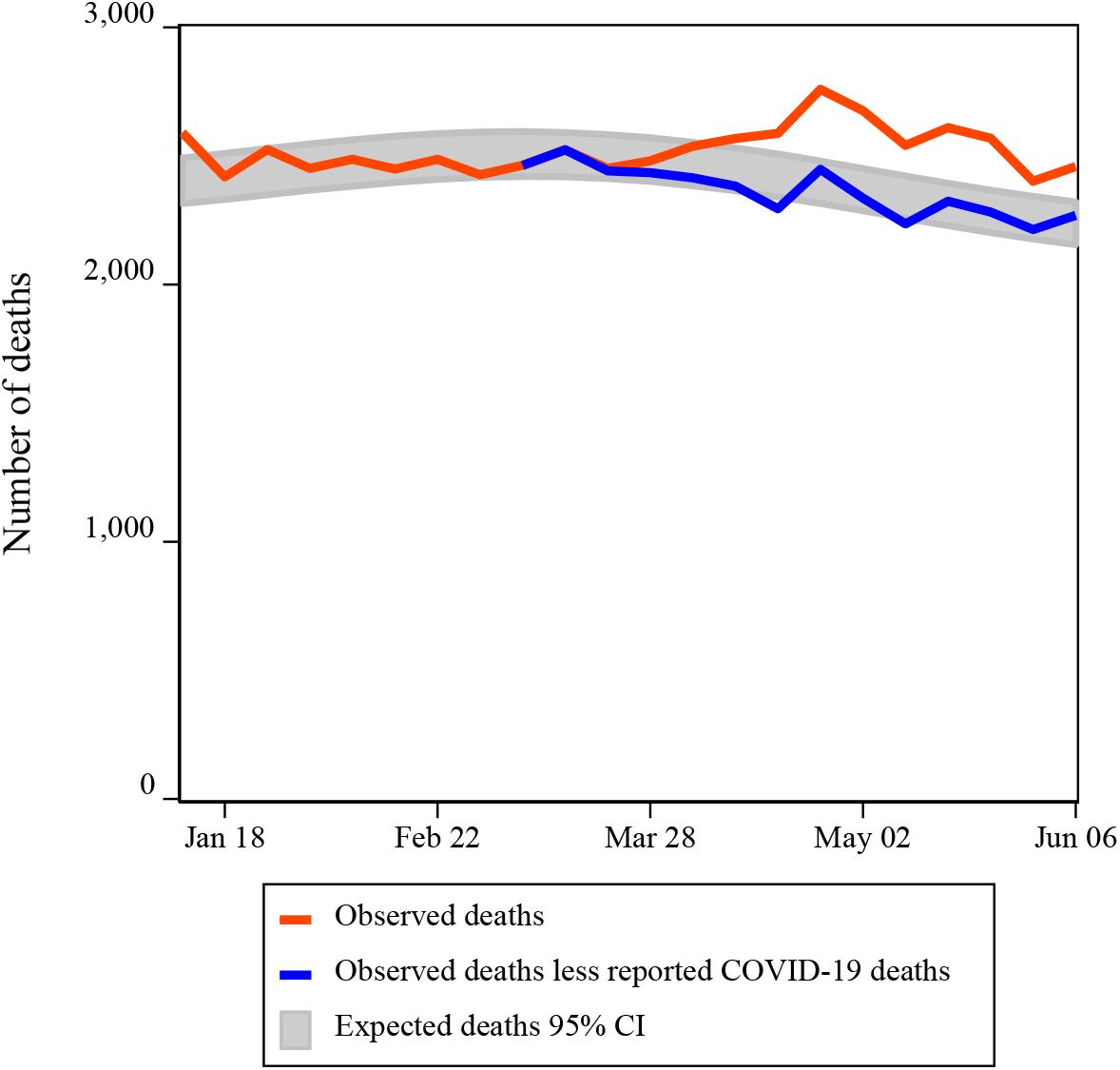
Excess deaths for all residents.

**Figure 2.**
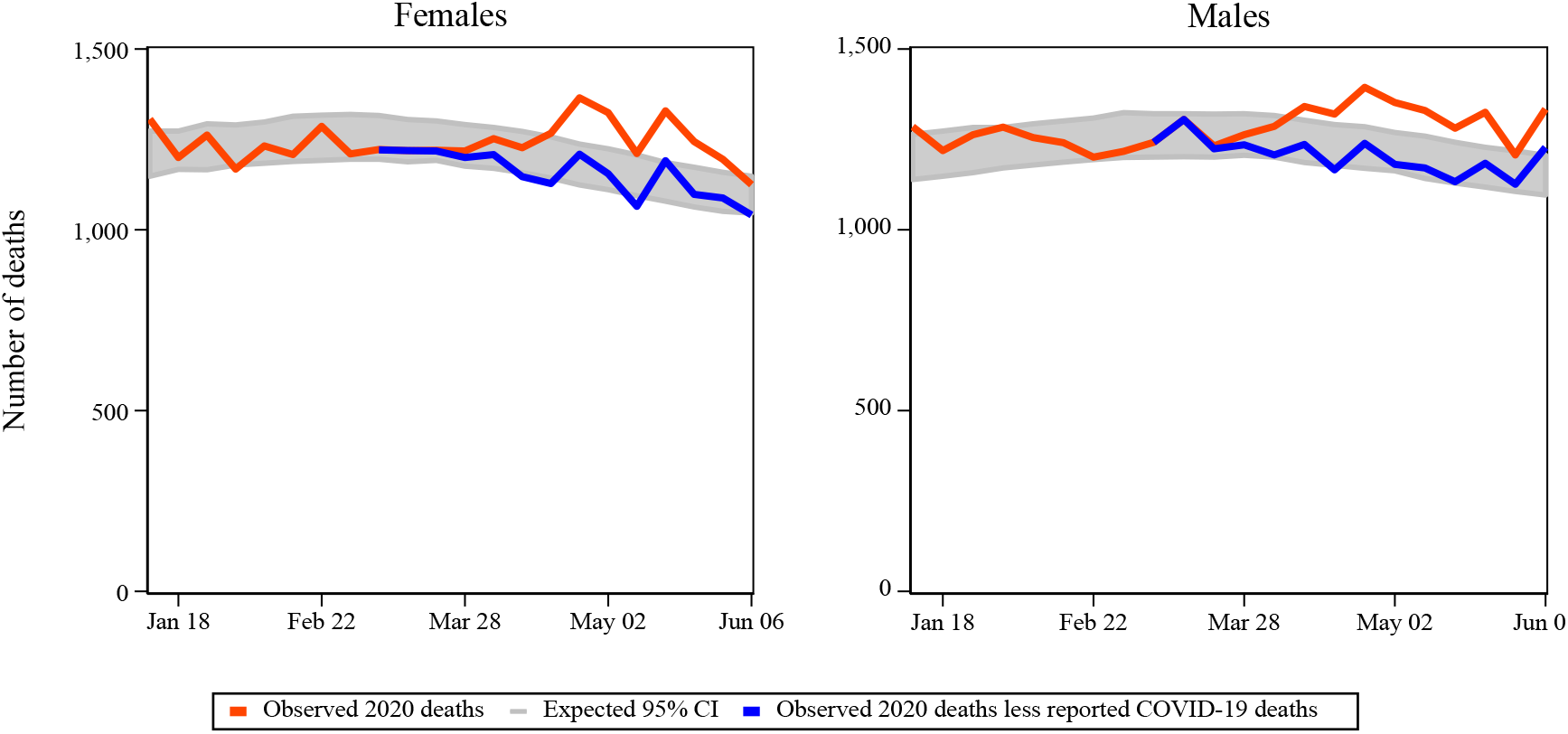
Excess deaths by gender.

**Figure 3.**
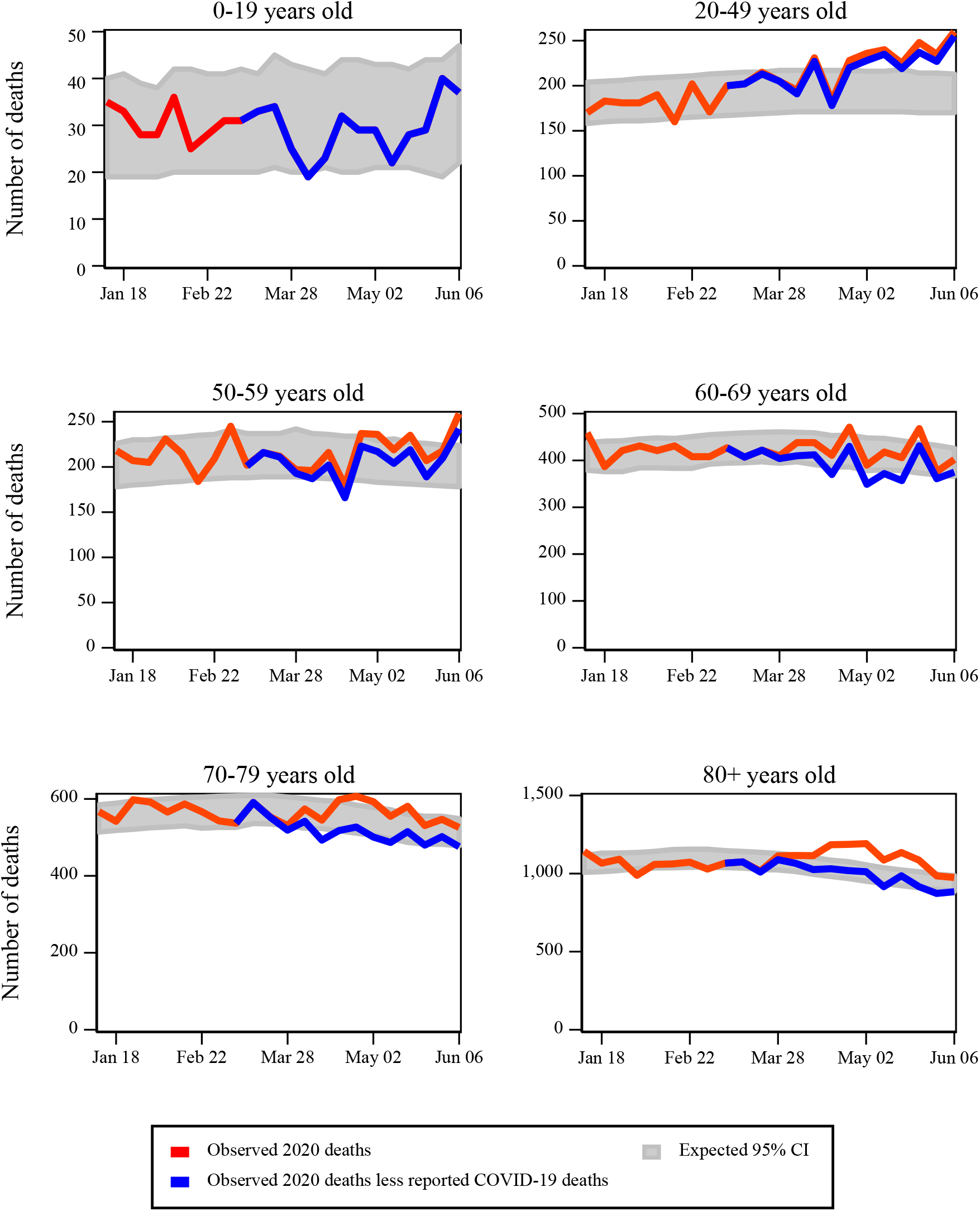
Excess deaths by age group.

**Figure 4.**
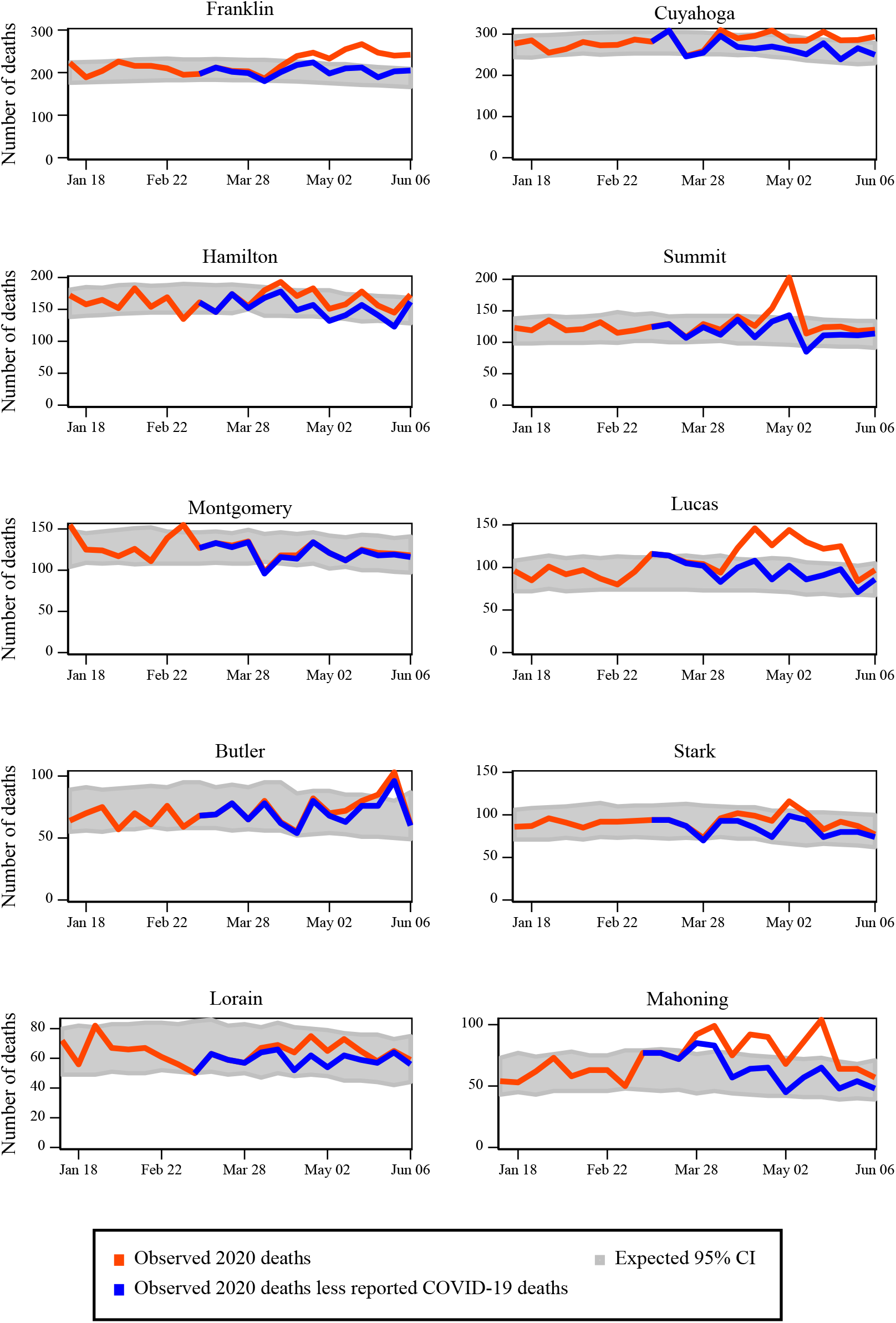
Excess deaths by county.

## Results

During our observation period the state of Ohio reported 2,564 COVID-19 deaths. Over those same weeks, there 30,643 all-cause deaths. For all residents, the number of excess all-cause deaths was 2,088 (95% CI 1,119-3,119) (figure 1). The reported number of COVID-19 deaths appears to largely explain the excess deaths across all residents, as the observed number of deaths less the reported number of COVID-19 deaths falls in the 95% confidence interval for the expected number of deaths. The number of excess deaths peaked in late April but the presence of excess deaths persisted throughout the observaton period.

Figure 2 shows for both females and males that the observed number of deaths exceeded the 95% confidence interval for expected deaths in the last eight weeks of the observation period. The patterns are relatively similar across genders except for an uptick for males during the week ending June 6, 2020.

The graphs by age group (Figure 3) show that the 80+ years old group had sustained observed deaths above the 95% prediction interval from the week ending April 11, 2020 through the week ending May 23, 2020. The 40-49 age group had observed deaths that exceeded the prediction interval from late April through the end of the observation period. Further, the number of observed deaths less the reported COVID-19 deaths was also above the prediction interval, suggesting that the reported number of COVID-19 deaths fell well short of explaining the number of excess deaths for that age group. For the entire observation period, the estimated number of excess deaths was 366 while the reported number of COVID-19 deaths was only 66. This finding is in contrast to the older age groups where the number of reported COVID-19 deaths exceeded the estimated number of excess deaths.

Figure 4 displays the estimates for the ten most populated counties in Ohio, listed in descending population order. There does not appear to be a clear association between population and the rate of excess deaths. The top three counties in terms of excess deaths are ranked first, sixth, and tenth in population. The most populated county (Franklin) had the most excess deaths (384; 95% CI 120, 674), while the number of excess deaths in the tenth-most populated (Mahoning) is nearly one-third of the observed deaths (31.8%; 95% CI 8.0, 47.2).

## Discussion

We analyzed administrative data and employed a rigorous methodology to provide insight into the difficult task of estimating the number of deaths caused by COVID-19. Our findings indicate potential disparities in the mortality effects of COVID-19, with excess mortality especially pronounced among those aged 80 and over and for those living in nursing home or long-term care facilities, while fewer than expected deaths were occurred at hospice facilities. We also found substantial levels of unexplained excess mortality for those in the 20-49 age group.

The number of reported COVID-19 deaths largely explained the levels of excess deaths that we estimated, but to varying degrees. For most of the groups for which reported COVID-19 deaths were available, the number of reported deaths exceeded the estimated number of excess deaths. While these findings stand in contrast with earlier work^9^ that found reported deaths to be less than the estimated number of excess deaths, our result may reflect that some COVID-19 deaths were not excess in the sense that the death would have occurred due to another cause if the individual had not contracted COVID-19. Also, fewer than expected deaths due to external causes (e.g., fewer vehicular fatalities due to stay-at-home behavior) may have reduced our estimates of excess deaths.

However, there were instances where the number of reported COVID-19 deaths was less than our excess death estimate, most notably for the 20-49 age group for which our estimate of excess deaths was six times the number of reported deaths. A key question is whether this discrepancy is due to unreported COVID-19 deaths for this age group, an increase in the number of deaths due to other causes, or some combination of the two. To attempt to gain insight into this discrepancy, Appendix Figure 1 shows the age-adjusted mortality rates for this age group by place of death for 2010-2019 and for 2020. The largest difference across the time periods is for deaths that occur at home. While this finding does not explain the far larger number of excess deaths than reported deaths, the higher mortality rate for deaths at home in 2020 may indicate that some individuals did not receive timely emergency care.

The substantial variation across counties highlights local variation of the COVID-19 outbreak. Not only do the counties with the highest number of excess deaths have varying population levels, but they are geographically distant from each other. Further, the trends in excess deaths differed across the three counties. While Lucas and Mahoning had greatly reduced the number of excess deaths by the end of the period, Franklin county had persistent levels over the last five months of the observation period. The extent to which the number of reported COVID-19 deaths explained the excess deaths also varied, where in Franklin county the number of reported COVID-19 deaths was 88% of the estimated excess deaths. In Lucas and Mahoning counties, the values were 75% and 72%, respectively.

Our study has several limitations. Our analysis was limited to one, albeit relatively large, U.S. state. There has been significant variation in the effects of COVID-19 across and within countries, so our findings do not necessarily apply universally. Our data were limited to the first several months of the outbreak and thus our results may not pertain to later stages. The methodology we employ does not definitively identify excess deaths as being due to COVID-19. Stressful events such as COVID-19 can lead to negative health outcomes such as increased incidence of cardiovascular events,^20^ poor medication adherence,^21^ and hypertension.^22^ Potential increases in mortality due to these types of effects would have been included in our estimates of excess deaths.

Our mortality data were imperfect in that the 2019 and 2020 data were provisional and not finalized. The mortality data were based on state and county of residence at the time of death, rather than the location of occurrence. This choice was made to align the data with the reported number of COVID-19 deaths, but insight could be gained by exploring potential differences from basing the data on where the death occurred. As noted in the Supplementary Appendix, the date of death had to be approximated for ten of the 2,564 deaths in the reported COVID-19 deaths.

The approach we employed indicated important differences in the effects of COVID-19 across demographic groups and identified potential shortcomings in published data. Our methodology can be applied in jurisdictions that provide timely access to death certificate data. While the results may not offer perfect insight into the precise causes of death, they can provide information to help craft effective policy and medical responses. Further, the methodology is robust to the imperfect coding of deaths that is common in crisis environments.

While our study is specific to Ohio, given the state’s large population and similarities with the U.S. as a whole, our results may improve the understanding of COVID-19 elsewhere. Policy makers, public health officials, and health care providers may be able to apply our findings to mitigate the destructive effects of COVID-19 and better prepare for future epidemics.

## Data Availability

The data and code employed in the analysis are available at https://github.com/troyquast/covid19_ohio.

## Acknowledgements

We thank the Ohio Department of Health for publishing timely mortality and COVID-19 data. The Department specifically disclaims responsibility for any analyses, interpretations or conclusions.

## Appendix

### Data sources

The Ohio Public Health Information warehouse is located at http://publicapps.odh.ohio.gov/EDW/DataCatalog. The data for the analyses were obtained on July 14, 2020.

The Ohio COVID-19 dashboard is located at https://coronavirus.ohio.gov/wps/portal/gov/covid-19/dashboards/overview. These data were obtained on July 14, 2020 and were limited to deaths that occurred through June 6, 2020. There were valid dates of death during the observation period for 2,571 deaths. Sixteen observations were listed with “Unknown” dates of death. Three of these observations had a valid hospital admission date. For these observations, the date of death was approximated as seven days after the admission date. Of these three observations, one had an approximated date that fell within the observation period and was thus included in our data. The thirteen remaining observations did not have a valid hospital admission date but did have a valid COVID-19 onset date value. For these observations, the date of death was approximated as fourteen days after the onset date. Of these ten observations, nine had an approximated date that fell within the observation period and was thus included in the sample. The final dataset contained 2,565 deaths, one of which had a date of death of March 1, 2020 and was thus not included in our observation period.

**Appendix Table 1.**
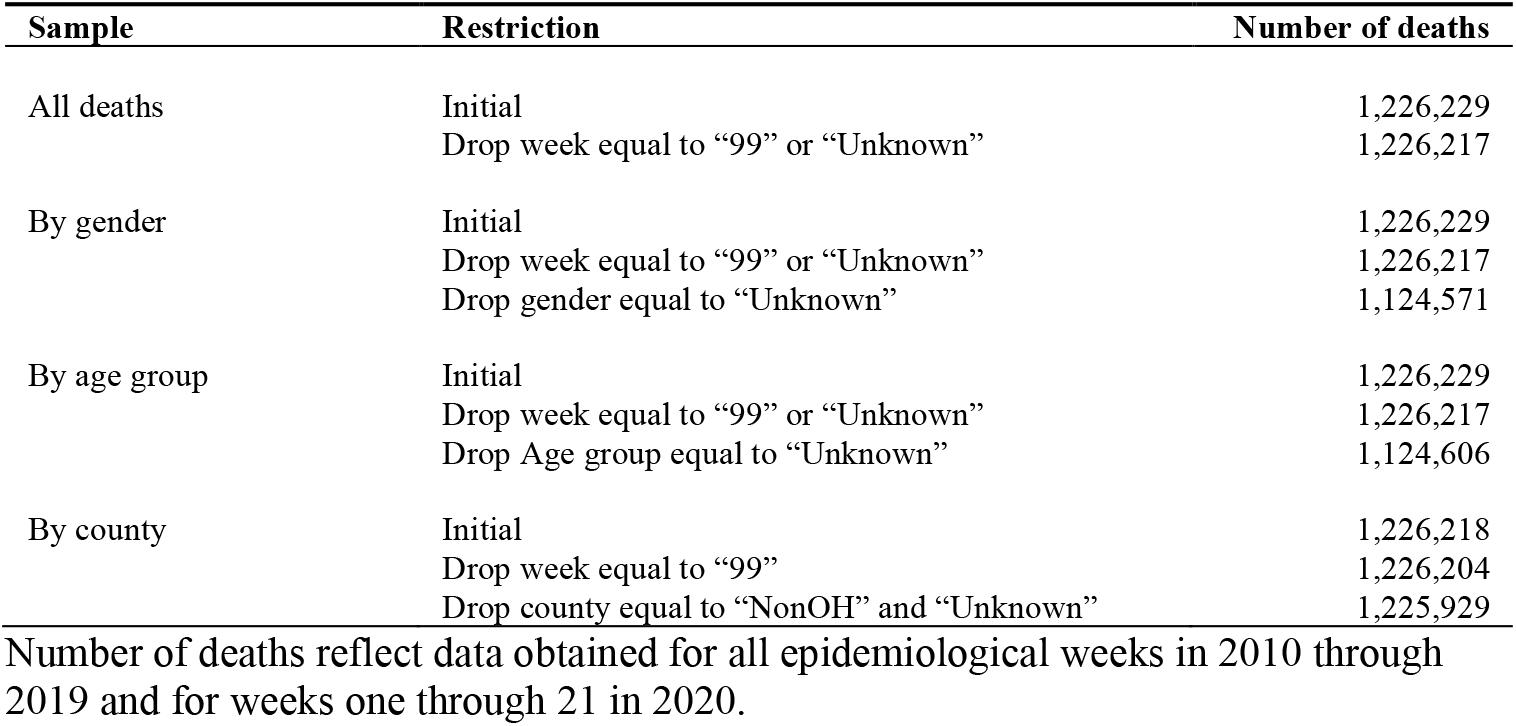
Number of deaths by sample after restricting to usable observations.

**Appendix Table 2.**
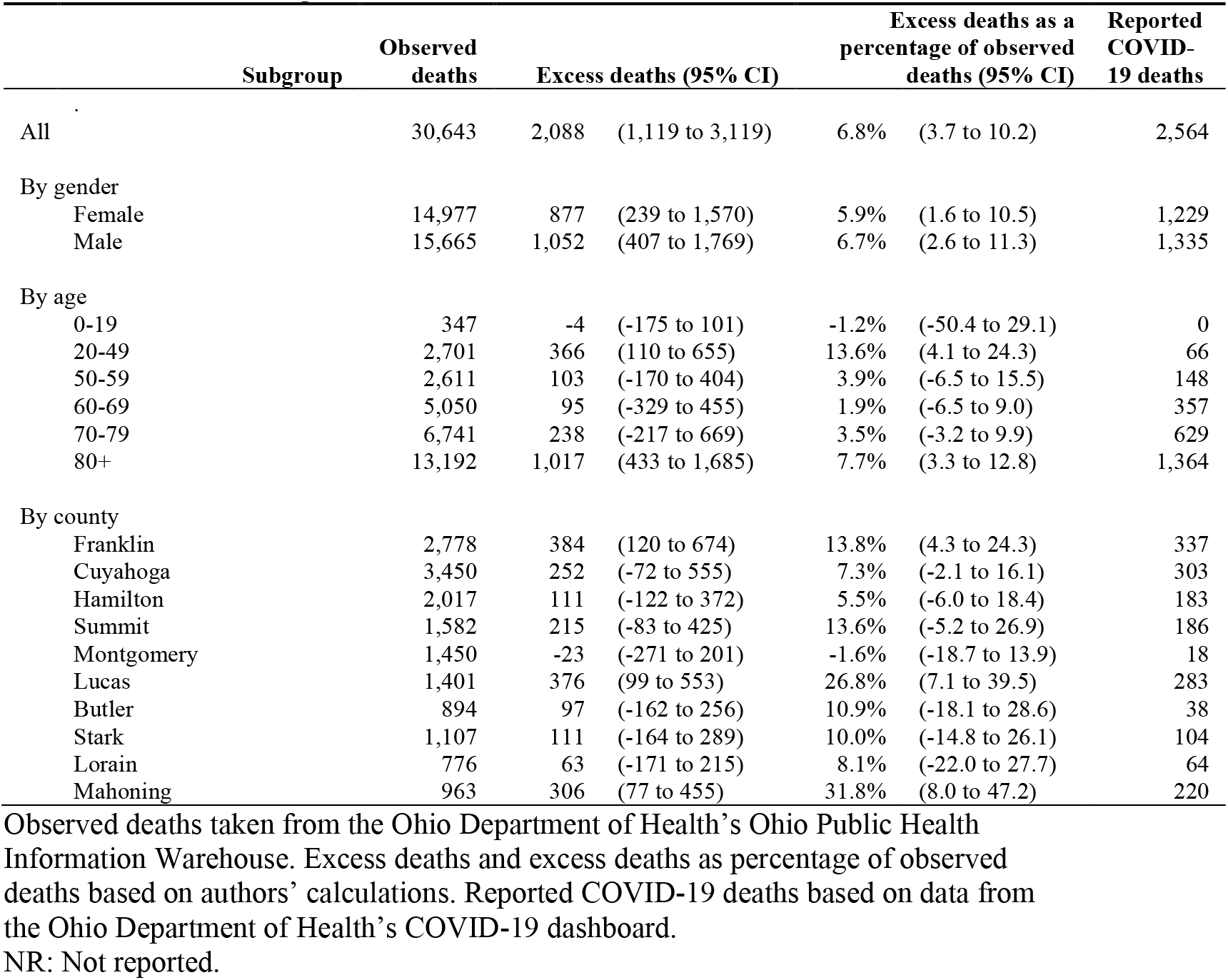
Observed and excess deaths and reported COVID-19 deaths in Ohio from March 15, 2020 through June 6, 2020.

| Subgroup | Observed deaths | Excess deaths (95% CI) | Excess deaths as a percentage of observed deaths (95% CI) | Reported COVID-19 deaths |
| --- | --- | --- | --- | --- |
| All | 30,643 | 2,088 (1,119 to 3,119) | 6.8% (3.7 to 10.2) | 2,564 |
| By gender |  |  |  |  |
| Female | 14,977 | 877 (239 to 1,570) | 5.9% (1.6 to 10.5) | 1,229 |
| Male | 15,665 | 1,052 (407 to 1,769) | 6.7% (2.6 to 11.3) | 1,335 |
| By age |  |  |  |  |
| 0-19 | 347 | -4 (-175 to 101) | -1.2% (-50.4 to 29.1) | 0 |
| 20-49 | 2,701 | 366 (110 to 655) | 13.6% (4.1 to 24.3) | 66 |
| 50-59 | 2,611 | 103 (-170 to 404) | 3.9% (-6.5 to 15.5) | 148 |
| 60-69 | 5,050 | 95 (-329 to 455) | 1.9% (-6.5 to 9.0) | 357 |
| 70-79 | 6,741 | 238 (-217 to 669) | 3.5% (-3.2 to 9.9) | 629 |
| 80+ | 13,192 | 1,017 (433 to 1,685) | 7.7% (3.3 to 12.8) | 1,364 |
| By county |  |  |  |  |
| Franklin | 2,778 | 384 (120 to 674) | 13.8% (4.3 to 24.3) | 337 |
| Cuyahoga | 3,450 | 252 (-72 to 555) | 7.3% (-2.1 to 16.1) | 303 |
| Hamilton | 2,017 | 111 (-122 to 372) | 5.5% (-6.0 to 18.4) | 183 |
| Summit | 1,582 | 215 (-83 to 425) | 13.6% (-5.2 to 26.9) | 186 |
| Montgomery | 1,450 | -23 (-271 to 201) | -1.6% (-18.7 to 13.9) | 18 |
| Lucas | 1,401 | 376 (99 to 553) | 26.8% (7.1 to 39.5) | 283 |
| Butler | 894 | 97 (-162 to 256) | 10.9% (-18.1 to 28.6) | 38 |
| Stark | 1,107 | 111 (-164 to 289) | 10.0% (-14.8 to 26.1) | 104 |
| Lorain | 776 | 63 (-171 to 215) | 8.1% (-22.0 to 27.7) | 64 |
| Mahoning | 963 | 306 (77 to 455) | 31.8% (8.0 to 47.2) | 220 |

Observed deaths taken from the Ohio Department of Health's Ohio Public Health Information Warehouse. Excess deaths and excess deaths as percentage of observed deaths based on authors' calculations. Reported COVID-19 deaths based on data from the Ohio Department of Health's COVID-19 dashboard. NR: Not reported.

**Appendix Figure 1.**
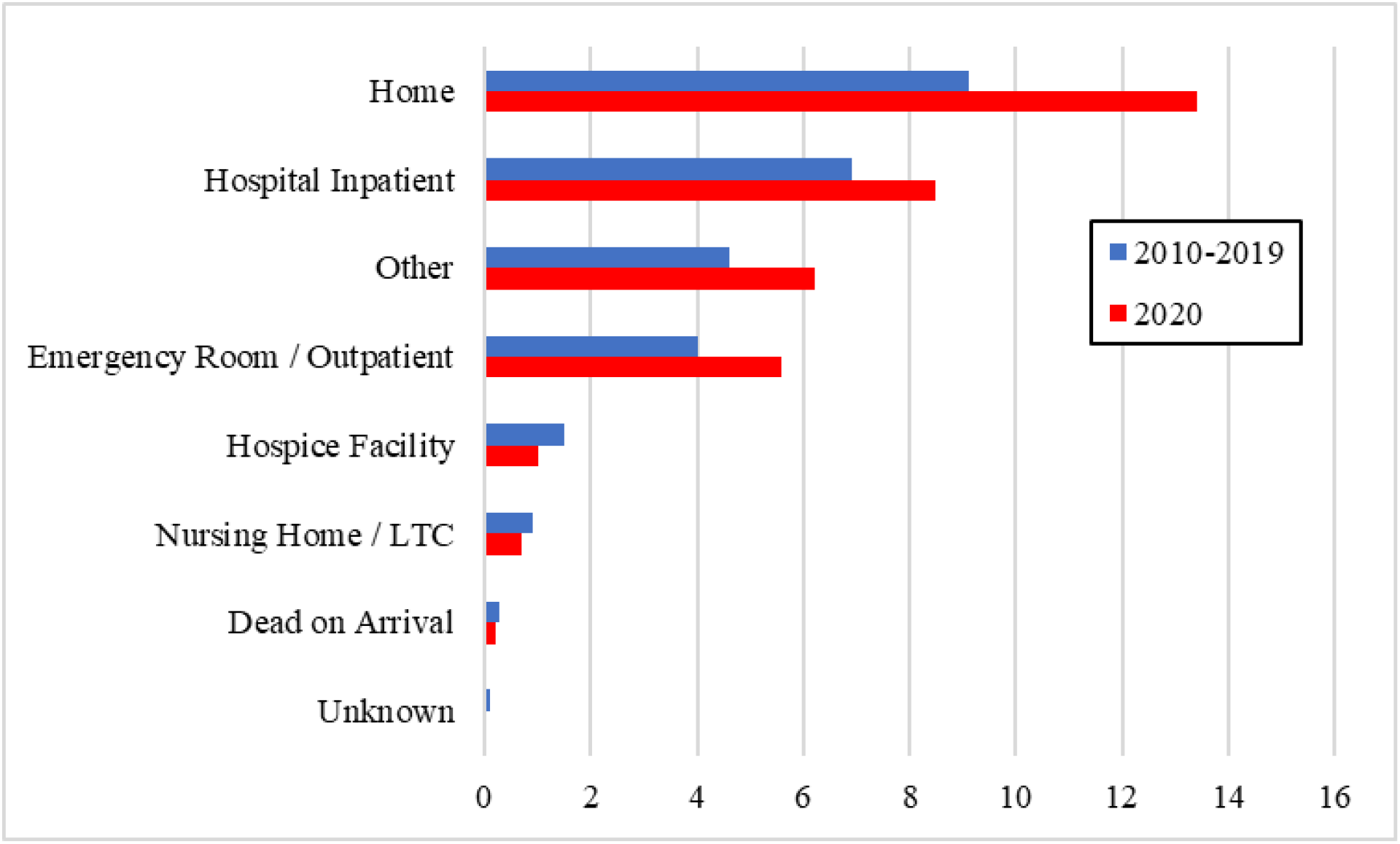
Age-adjusted mortality rates per 100,000 persons for 20-49 years old by place of death, 2010-2019 and 2020.

## References

1 Council of State & Territorial Epidemiologists. Position Statement: Standardized surveillance case definition and national notification for 2019 novel coronavirus disease (COVID-19). April 5, 2020. https://cdn.ymaws.com/www.cste.org/resource/resmgr/2020ps/Interim-20-ID-01_COVID-19.pdf (accessed May 23, 2020).

2 Resnick B, Scott D. America’s shamefully slow coronavirus testing threatens all of us. Vox. March 12, 2020. https://www.vox.com/science-and-health/2020/3/12/21175034/coronavirus-covid-19-testing-usa (accessed June 22, 2020).

3 Goh KJ, Wong J, Tien J-CC, et al. Preparing your intensive care unit for the COVID-19 pandemic: practical considerations and strategies. Critical Care 2020; 24:215.

4 Stephens KU, Grew D, Chin K, et al. Excess Mortality in the Aftermath of Hurricane Katrina: A Preliminary Report. Disaster Med Public Health Prep 2007; 1:15–20.

5 Santos-Burgoa C, Sandberg J, et al. Differential and persistent risk of excess mortality from Hurricane Maria in Puerto Rico: a time-series analysis. Lancet Planet Health 2018; 2:e478–e488.

6 Santos-Lozada AR, Howard JT. Use of Death Counts From Vital Statistics to Calculate Excess Deaths in Puerto Rico Following Hurricane Maria. JAMA 2018; 320:1491–1493.

7 Whitman S, Good G, Donoghue ER, Benbow N, Shou W, Mou S. Mortality in Chicago attributed to the July 1995 heat wave. Am J Public Health 1997; 87:1515–1518.

8 Krieger N, Chen JT, Waterman PD. Excess mortality in men and women in Massachusetts during the COVID-19 pandemic. Lancet 2020; 395:1829.

9 Weinberger D, Cohen T, Crawford F, et al. Estimating the early death toll of COVID-19 in the United States. medRXiv 2020; published online April 29. DOI:10.1101/2020.04.15.20066431 (preprint).

10 US Centers for Disease Control and Prevention. Excess Deaths Associated with COVID-19. June 17, 2020. https://www.cdc.gov/nchs/nvss/vsrr/covid19/excess_deaths.htm (accessed June 21, 2020).

11 Freitas ARR, Medeiros NM, Frutuoso L, et al. Use of excess mortality associated with the COVID-19 epidemic as an epidemiological surveillance strategy - preliminary results of the evaluation of six Brazilian capitals. medRxiv 2020; published online May 12. DOI: 10.1101/2020.05.08.20093617 (preprint).

12 Vieira A, Peixoto VR, Aguiar P, Abrantes A. Rapid estimation of excess mortality in times of COVID-19 in Portugal - Beyond reported deaths. medRxiv 2020; published online May 19. DOI: 10.1101/2020.05.14.20100909 (preprint).

13 Kontopantelis E, Mamas MA, Deanfield J, et al. Excess mortality in England and Wales during the first wave of the COVID-19 pandemic. medRxiv 2020; published online June 18. DOI: 10.1101/2020.05.26.20113357 (preprint).

14 McLeay S. Measuring Excess Mortality: A Second Look at Germany. Social Science Research Network 2020; published online May 27. DOI: 10.2139/ssrn.3626618 (preprint).

15 Modi C, Boehm V, Ferraro S, et al. How deadly is COVID-19? A rigorous analysis of excess mortality and age-dependent fatality rates in Italy. medRxiv 2020; published online May 14. DOI: 10.1101/2020.04.15.20067074 (preprint).

16 US Census Bureau. QuickFacts: Ohio; United States. https://www.census.gov/quickfacts/fact/table/OH,US/ PST045219 (accessed June 9, 2020)

17 Meckler L. Seven states, D.C. order all schools closed in effort to prevent spread of covid-19. Washington Post, March 13, 2020. https://www.washingtonpost.com/local/education/ohio-maryland-order-all-schools-closed-in-effort-to-prevent-spread-of-covid-19/2020/03/12/e4078b3a-6499-11ea-845d-e35b0234b136_story.html (accessed June 21, 2020).

18 O’Brien RD, Bauerlein V. How Coronavirus Remade American Life in One Weekend. Wall Street Journal, March 15, 2020. https://www.wsj.com/articles/coronavirus-remakes-american-life-in-a-weekend-11584293065 (accessed June 20, 2020).

19 Borchardt J. Ohio started reopening six weeks ago, hasn’t seen a coronavirus case surge. USA TODAY, June 18, 2020. https://www.usatoday.com/story/news/health/2020/06/18/ohio-reopening-one-month-later-no-surge-in-coronavirus-cases/3208493001/ (accessed June 20, 2020).

20 Steptoe A, Kivimäki M. Stress and cardiovascular disease. Nat Rev Cardiol. 2012; 9:360–370.

21 Kretchy IA, Owusu-Daaku FT, Danquah SA. Mental health in hypertension: assessing symptoms of anxiety, depression and stress on anti-hypertensive medication adherence. Int J Ment Health Syst. 2014; 8:25.

22 Sparrenberger F, Cichelero FT, Ascoli AM, et al. Does psychosocial stress cause hypertension? A systematic review of observational studies. J Hum Hypertens. 2009; 23:12–19.

